# A Hybrid CNN-Transformer Deep Learning Model for Differentiating Benign and Malignant Breast Tumors Using Multi-View Ultrasound Images

**DOI:** 10.1101/2025.08.24.25334030

**Authors:** Zhang Qi, Ruizhuo Li, Tang Pan, Zhang Jianxing, Guangxu Yang, Ziyi Lin, Mengjian Zhang, Chen Miao

## Abstract

Breast cancer is a leading malignancy threatening women’s health globally, making early and accurate diagnosis crucial. Ultrasound is a key screening and diagnostic tool due to its non-invasive, real-time, and cost-effective nature. However, its diagnostic accuracy is highly dependent on operator experience, and conventional single-image analysis often fails to capture the comprehensive features of a lesion. This study introduces a computer-aided diagnosis (CAD) system that emulates a clinician’s multi-view diagnostic process. We developed a novel hybrid deep learning model that integrates a Convolutional Neural Network (CNN) with a Transformer architecture. The model uses a pretrained EfficientNetV2 to extract spatial features from multiple, unordered ultrasound images of a single lesion. These features are then processed by a Transformer encoder, whose self-attention mechanism globally models and fuses their intrinsic correlations. To prevent data leakage, a strict lesion-level data partitioning strategy ensured a rigorous evaluation. On an internal test set, our hybrid CNN-Transformer achieved an accuracy of 0.960, a sensitivity of 0.967, a specificity of 0.954, and an Area Under the Curve (AUC) of 0.9788. On an external dataset, it demonstrated an accuracy of 0.940, a sensitivity of 0.952, a specificity of 0.929, and an AUC of 0.9730. Furthermore, in a prospective validation on a newly collected independent dataset, the model maintained robust performance with an accuracy of 0.952 and an AUC of 0.9801. These results significantly outperform those of a baseline single-image model, which achieved accuracies of 0.88 and 0.89 and AUC of 0.95 and 0.94 on the internal and external dataset, respectively. This study demonstrates that combining CNN with Transformers yields a highly accurate and robust diagnostic system for breast ultrasound. By effectively fusing multi-view information, our model aligns with clinical logic and shows potential for improving diagnostic reliability.

## 1. Introduction

Breast cancer stands as the most frequently diagnosed malignancy among women worldwide, posing a significant threat to their lives and health(1). Clinical evidence indicates that early detection, diagnosis, and treatment are paramount for improving patient prognosis and achieving higher five-year survival rates(2). Among the various imaging modalities, breast ultrasound has emerged as an pivotal first-line tool for the screening, diagnosis, and follow-up of breast diseases, owing to its unique advantages of being non-invasive, radiation-free, highly repeatable, real-time, and cost-effective(3).

However, the traditional ultrasound diagnostic process has notable limitations. The accuracy of a diagnosis is heavily reliant on the sonographer’s expertise, technical skill, and subjective judgment, which can lead to inter-observer variability and diagnostic discrepancies(4). In clinical practice, an experienced physician does not rely on a single, static image to assess a suspicious lesion. Instead, they perform a scan from multiple angles and planes to comprehensively observe multi-dimensional features such as the lesion’s margins, shape, internal echo pattern, and posterior acoustic characteristics(5). This process allows them to build a holistic, three-dimensional impression of the lesion before arriving at a final diagnosis. This integrative analysis is central to diagnostic accuracy, and a key challenge in intelligent medical imaging is to develop artificial intelligence (AI) that can effectively simulate and enhance this complex cognitive process.

In recent years, AI technologies, particularly deep learning (DL), have made transformative breakthroughs in medical image analysis(6, 7). Convolutional Neural Networks (CNNs), with their powerful ability to automatically learn hierarchical feature representations, have become the gold standard for processing grid-like data such as images(8–10). In the context of breast cancer diagnosis, various classic CNN architectures like VGG and ResNet have been successfully applied to classification tasks using mammography, histopathology, and ultrasound images, demonstrating performance comparable to, or even superior to, that of human experts on specific datasets(11–15). These models can autonomously learn subtle textural, morphological, and boundary features associated with malignancy, reducing the reliance on manually engineered features and paving the way for objective, efficient, and automated diagnosis.

Despite the remarkable success of CNN-based models, the majority of existing studies treat each 2D ultrasound image as an independent sample(16, 17). This methodology tends to diverge from standard clinical workflows, where diagnosis relies on the cumulative evidence derived from a sequence of images rather than isolated frames. Unlike single-image models that analyze specific views in isolation, clinical reasoning involves determining how collective findings contribute to a final assessment. Addressing this disparity—by evolving from static, single-view analysis to holistic, patient-level integration—represents a significant avenue for advancing intelligent medical imaging. This study is motivated by the need to bridge this gap, positing that the intelligent analysis of breast ultrasound should treat the complete set of multi-view images from a single lesion as a holistic, information-rich sequence.

To model image sequences, some researchers have explored hybrid architectures, such as combining a CNN with a Recurrent Neural Network (RNN) or its variant, Long Short-Term Memory (LSTM)(18). While valuable for data with a clear temporal or spatial order, these models are ill-suited for the unordered sets of images typical in a breast ultrasound examination. The arbitrary acquisition order of these images is less compatible with the inherent sequential dependency of RNN. To overcome this, we introduce the Transformer model, originally developed for natural language processing(19, 20). The Transformer’s core self-attention mechanism allows it to weigh the importance of all elements in a sequence simultaneously, regardless of their position. This property, known as permutation invariance, closely aligns with the clinical need to synthesize information from a collection of unordered images. Based on this analysis, this study proposes a new intelligent diagnostic paradigm that more closely mirrors clinical logic. The primary contributions are:

- We developed an end-to-end hybrid model combining EfficientNetV2 and a Transformer for breast ultrasound classification.
- We innovatively treats multiple images originating from a single tumor as a sequence, leveraging the Transformer’s global relationship modeling capabilities for comprehensive analysis.
- Rigorous, tumor-level experiments demonstrated the superior performance (an AUC of 0.9730 on an external dataset) of the model, which surpassed traditional single-image models.
- The solution offers a computationally effective approach to the core clinical challenge of multi-view information integration, and it indicates potential for future high-precision, reliable Computer-Aided Diagnosis (CAD) systems.

## 2. Materials and Methods

This section provides a detailed description of the data sources, preprocessing pipeline, model architecture, experimental configuration, and performance evaluation standards to ensure the study’s reproducibility. The overall workflow, illustrated in Fig 1, comprises three main stages: data preparation, model construction and training, and performance evaluation.

**Fig 1:**
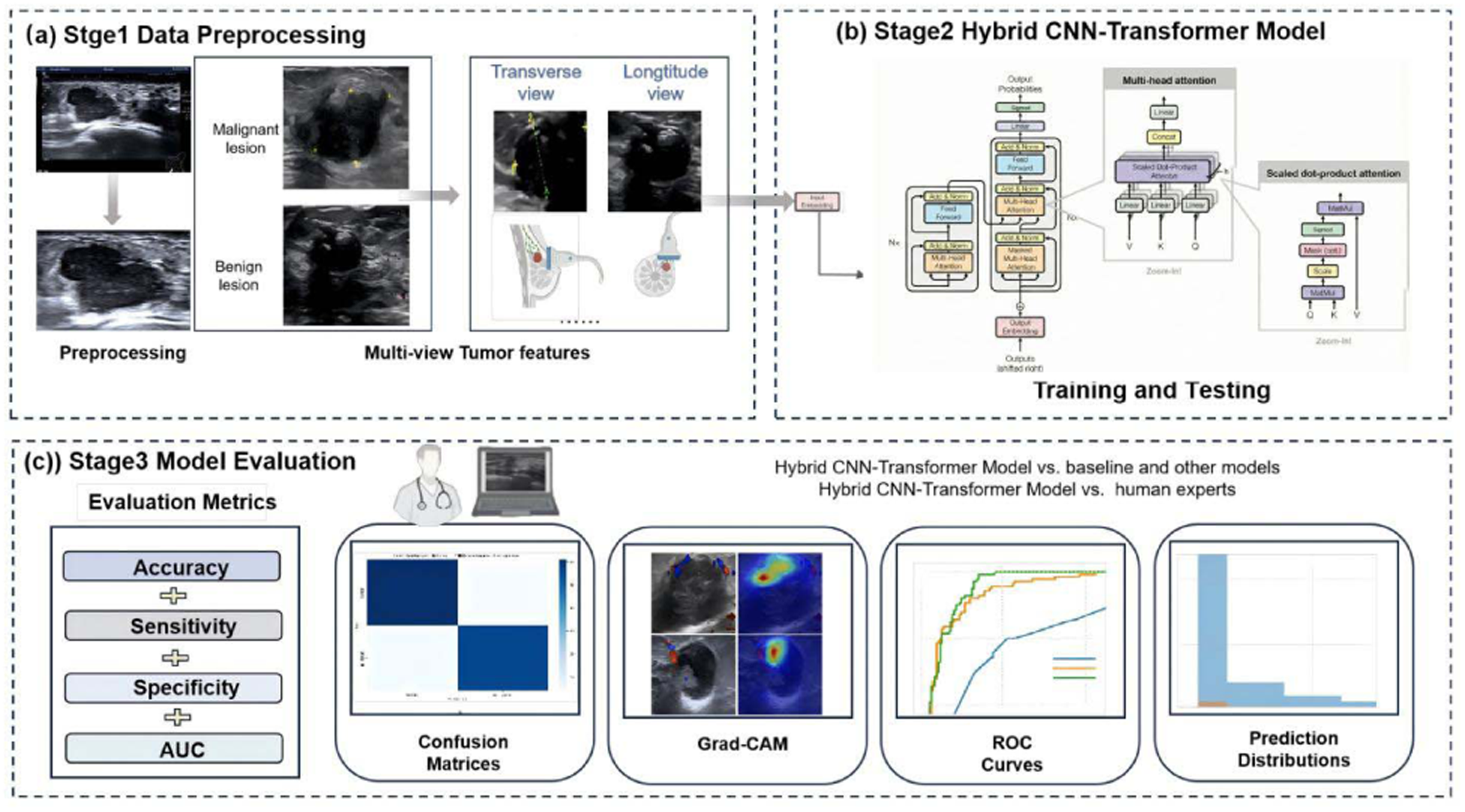
overall workflow.

### 2.1. Study Population and Ethical Approval

This retrospective study was conducted in accordance with the Declaration of Helsinki and approved by the Institutional Ethics Committee of The Second Affiliated Hospital of Guangzhou University of Chinese Medicine (Approval No. ZE2023-427, 05/12/2023). The requirement for informed consent was waived due to the retrospective nature of the study.

The case inclusion process is illustrated in Fig 2. We assessed medical records from two independent sources. The primary dataset was derived from The Second Affiliated Hospital of Guangzhou University of Chinese Medicine between June 2023 and April 2025. Inclusion criteria were: (1) patients with complete clinical and final pathological results who underwent preoperative ultrasound; (2) preoperative ultrasound with transverse or sagittal views and high-quality images; (3) patients who had not received neoadjuvant chemotherapy or radiotherapy. Exclusion criteria were: (1) poor ultrasound image quality; (2) an interval between ultrasound and pathology greater than two weeks; (3) prior surgery, biopsy, or chemotherapy. Out of 980 initially screened patients, 140 were excluded. The final primary dataset comprised 840 patients(age range: 11–88 years; mean age: 51.28 ± 13.55 years), consisting of 430 benign patients (1566 images) and 410 malignant patients (2220 images).

**Fig 2:**
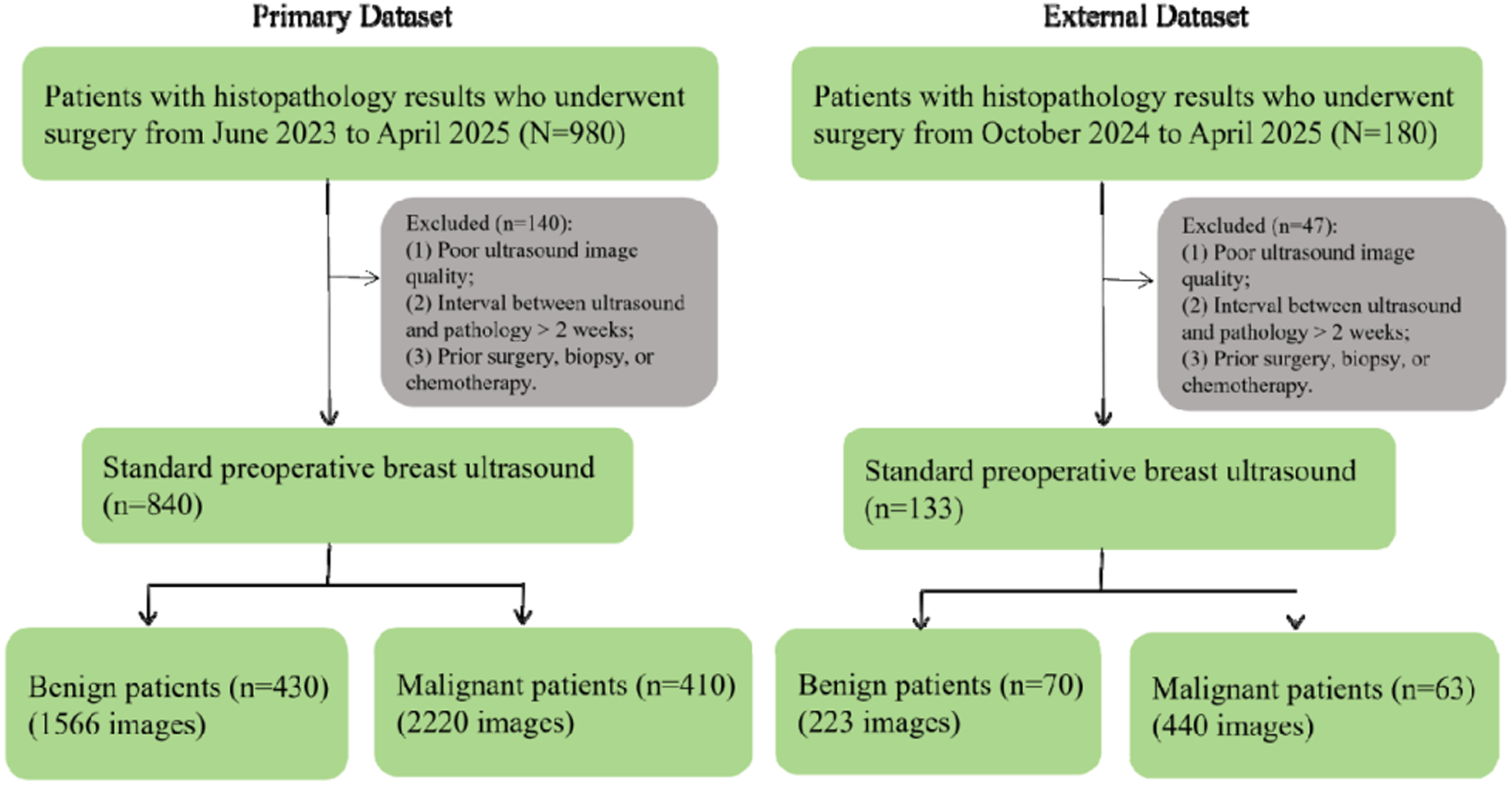
The case inclusion Process.

The external dataset was collected from the The Affiliated Hospital of Zunyi Medical University from October 2024 to April 2025. Following the same exclusion criteria, 47 out of 180 screened patients were excluded. The final external dataset included 133 patients, comprising 70 benign patients (223 images) and 63 malignant patients (440 images).

For model development, the datasets were categorized into training, validation, test, and external sets. Detailed clinical characteristics, including patient age distribution and clinical tumor size ( T1≤2.0cm and T2 2.1–5.0cm), are summarized in Table 1.

**Table 1:**
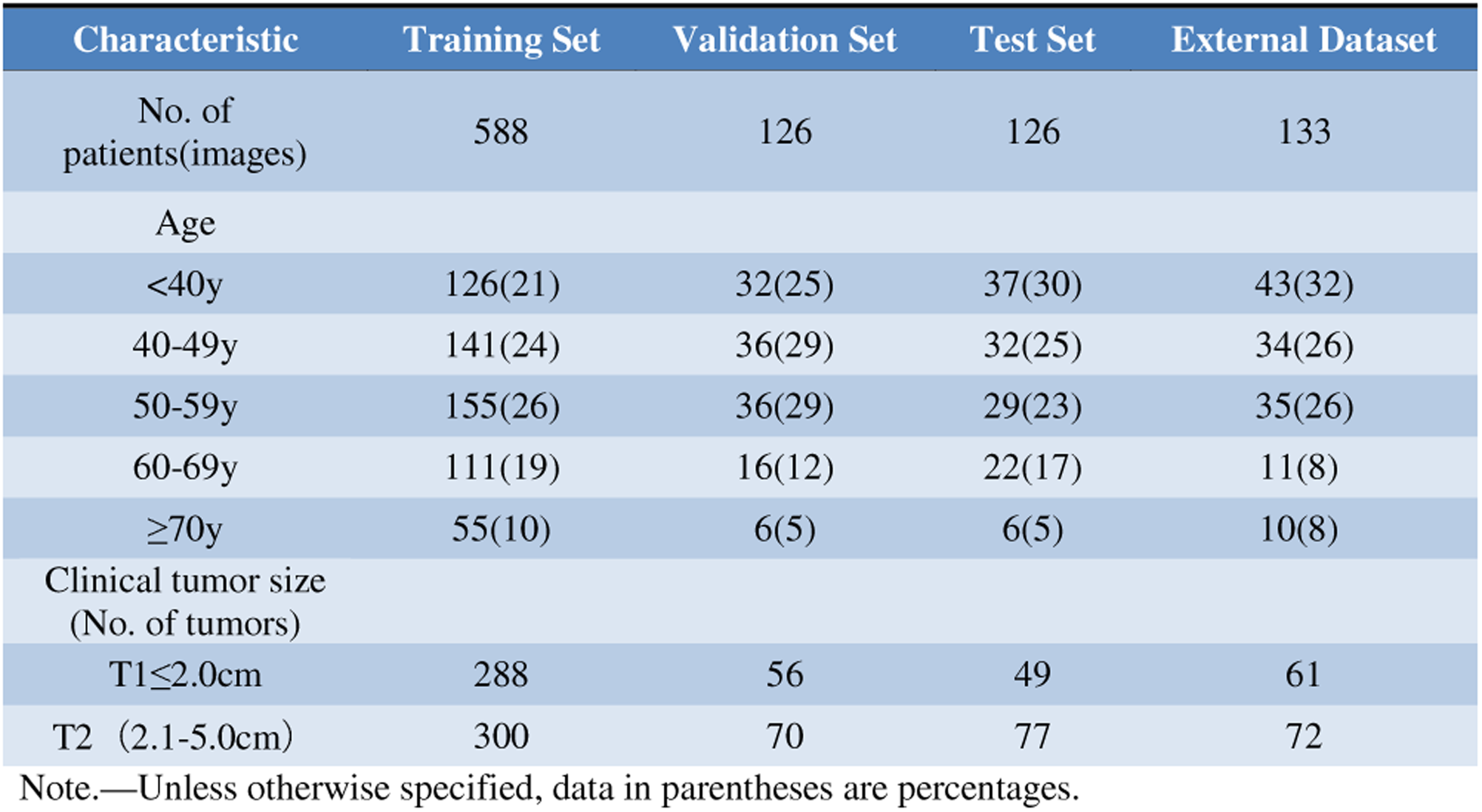
The clinical characteristics of all patients.

### 2.2. Data Acquisition and Preprocessing

Ultrasound examinations were performed by experienced physicians using GE LOGIQ E9 and Philips EPIQ 7C systems with high-frequency linear probes (7.5–13.0 MHz). Standard images of each lesion were acquired and stored in the picture archiving and communication system (PACS). The raw DICOM images were converted to JPG format. Two physicians, each with over five years of experience in breast ultrasound diagnosis, manually cropped the images to isolate the lesion and remove extraneous information, such as patient data and measurement markers in Fig 3. This step ensures the model focuses on relevant pathological features. All images were then resized to 300×300 pixels to match the model’s input dimensions, and pixel values were normalized to a range of [0, 1].

**Fig 3.**
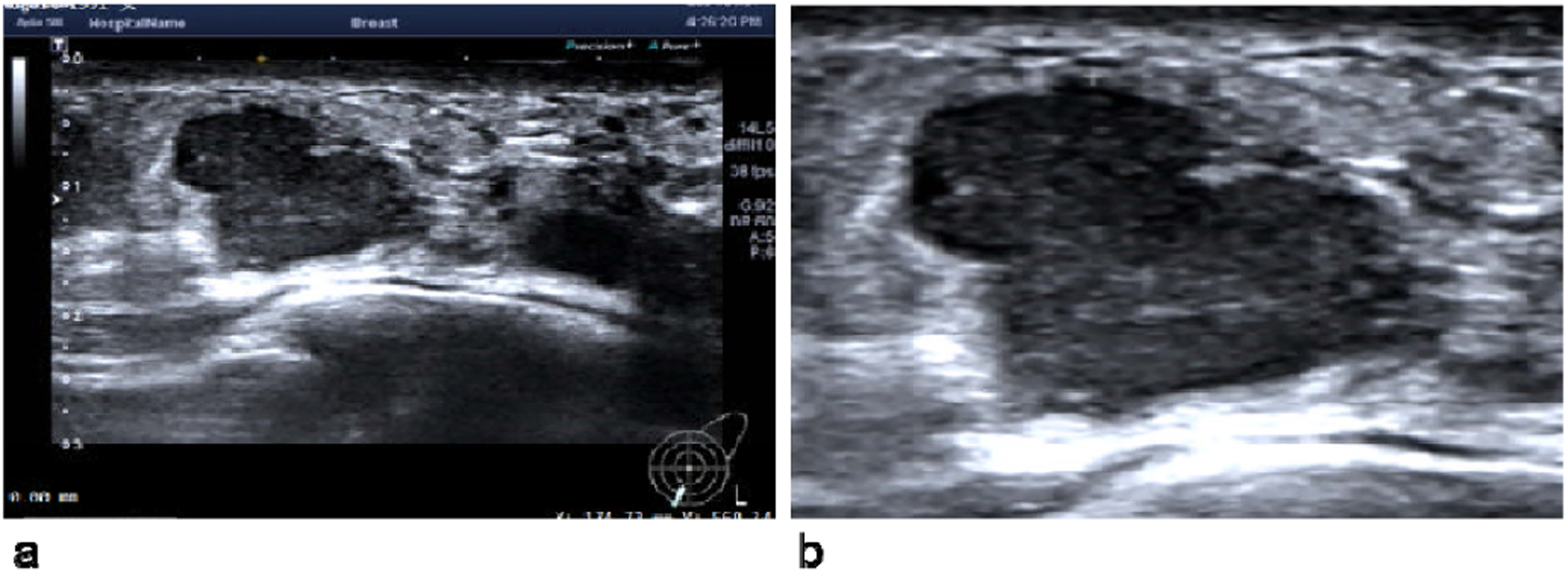
Pre- and Post-processing of Ultrasound Images; Example of the image preparation workflow. (a) The original ultrasound image as exported from the PACS, containing patient information and measurement markers. (b) The image after being manually cropped by an expert physician to isolate the lesion of interest for model input.

### 2.3. Dataset Partitioning Strategy

A scientifically rigorous dataset partitioning strategy is fundamental to the objective and reliable evaluation of AI models in medical imaging. A common pitfall is the random allocation of multiple images from the same patient’s tumor into the training, validation, and test sets. This practice leads to severe data leakage, as the model may learn to identify patient-specific artifacts rather than generalizable pathological features. This results in an artificially inflated performance on the test set, which plummets when the model is exposed to new data in a real-world setting. To address this issue, this study employed a strict per-lesion partitioning methodology. Specifically, all images associated with a single tumor were treated as an indivisible unit and assigned exclusively to one of the three sets: training, validation, or test. The internal dataset was partitioned using a 70%:15%:15% ratio, ensuring complete independence at the lesion level. The entire external dataset was held out as a separate, independent test set to evaluate the model’s generalization performance. The data distribution is detailed in Table 2.

**Table 2:** Dataset distribution for the baseline and hybrid models.

| Model Type | Data Unit | Training Set | Validation Set | Test Set | External Dataset | Total |
| --- | --- | --- | --- | --- | --- | --- |
| Baseline | Images | 2657 | 590 | 539 | 663 | 4449 |
| Hybrid | Tumors | 588 | 126 | 126 | 133 | 973 |

### 2.4. Handling Multi-View Data

A critical challenge in multi-view learning is the variable number of images available per lesion (ranging from 2 to 10). We implemented a robust technical strategy to handle this variability:

#### 2.4.1. Fixed Max Length and Padding

We defined a hyperparameter max_images_per_lesion = 10.

- Truncation: If a lesion has >10 images, only the first 10 are used.
- Zero-Padding: If a lesion has *N* < 10 images, we generate 10 - *N* dummy images consisting of all-zero tensors. This ensures that the input tensor to the model always has a fixed shape.

#### 2.5.1. Baseline Model: EfficientNetV2 Single-Image Classifier

The baseline model was implemented using a standard transfer learning paradigm to establish a performance benchmark for a conventional single-image approach.

- **Backbone:** The efficientnet_v2_s model, pre-trained on the ImageNet dataset, was selected as the feature extractor due to its excellent balance between performance and efficiency.
- **Feature Extraction:** The parameters of the convolutional layers in the backbone were frozen, allowing it to function as a fixed feature extraction tool.
- **Classification Head:** The original fully connected layer was replaced with a custom head consisting of a linear layer (1280 input dimensions) and a single output neuron with a sigmoid activation function to produce a binary probability of malignancy.

#### 2.4.2. Attention Masking

To ensure the zero-padding does not influence the diagnosis, we generated a binary Attention Mask for each sample. The mask contains 1 for real images and 0 for padded images. In the Transformer’s self-attention calculation, the similarity scores corresponding to padded positions are set to negative infinity before the Softmax operation. This forces the attention weights for padded images to become effectively zero, meaning they contribute nothing to the feature aggregation.

#### 2.4.3. Classification Token Aggregation

Instead of simple averaging, which dilutes strong signals, we prepend a learnable token to the sequence of image features. The Transformer learns to update this token with information aggregated from all valid views. The final classification is performed solely on the state of this token.

### 2.5. Model Architectures

This study designed and compared two distinct models: a single-image classification baseline and the proposed hybrid CNN-Transformer model.

#### 2.5.2. Proposed Hybrid Model

This model consists of two stages:

- **Local Feature Extraction:** Each of the *N* images in a patient’s lesion is passed independently through the EfficientNetV2 backbone, resulting in a sequence of feature vectors *F*={*f*_1_,*f*_2_,…,*f_N_*}, where *f_i_* □□^1280^.
- **Global Feature Fusion:** The sequence was fed into a Transformer Encoder.

- Hyper parameter: To ensure optimal performance, we conducted a systematic grid search for hyperparameter optimization. We evaluated dropout rates of {0.0,0.1,0.2,0.3,0.4,0.5}, attention heads of {4,8,16}, and network depths of {4,6,8,12,16} layers. Based on the validation set performance of AUC, the final model was configured with 8 layers, 8 attention heads, and a dropout rate of 0.1 in Fig 4.
- **Positional Encoding:** Crucially, we removed the positional encoding typically used in NLP Transformers. This enforces permutation invariance, ensuring the model treats the input as an unordered set rather than a sequence.
- **Output:** The final embedding of the token is passed to a Multi-Layer Perceptron (MLP) head with a Sigmoid activation to predict the probability of malignancy.

**Fig 4.**
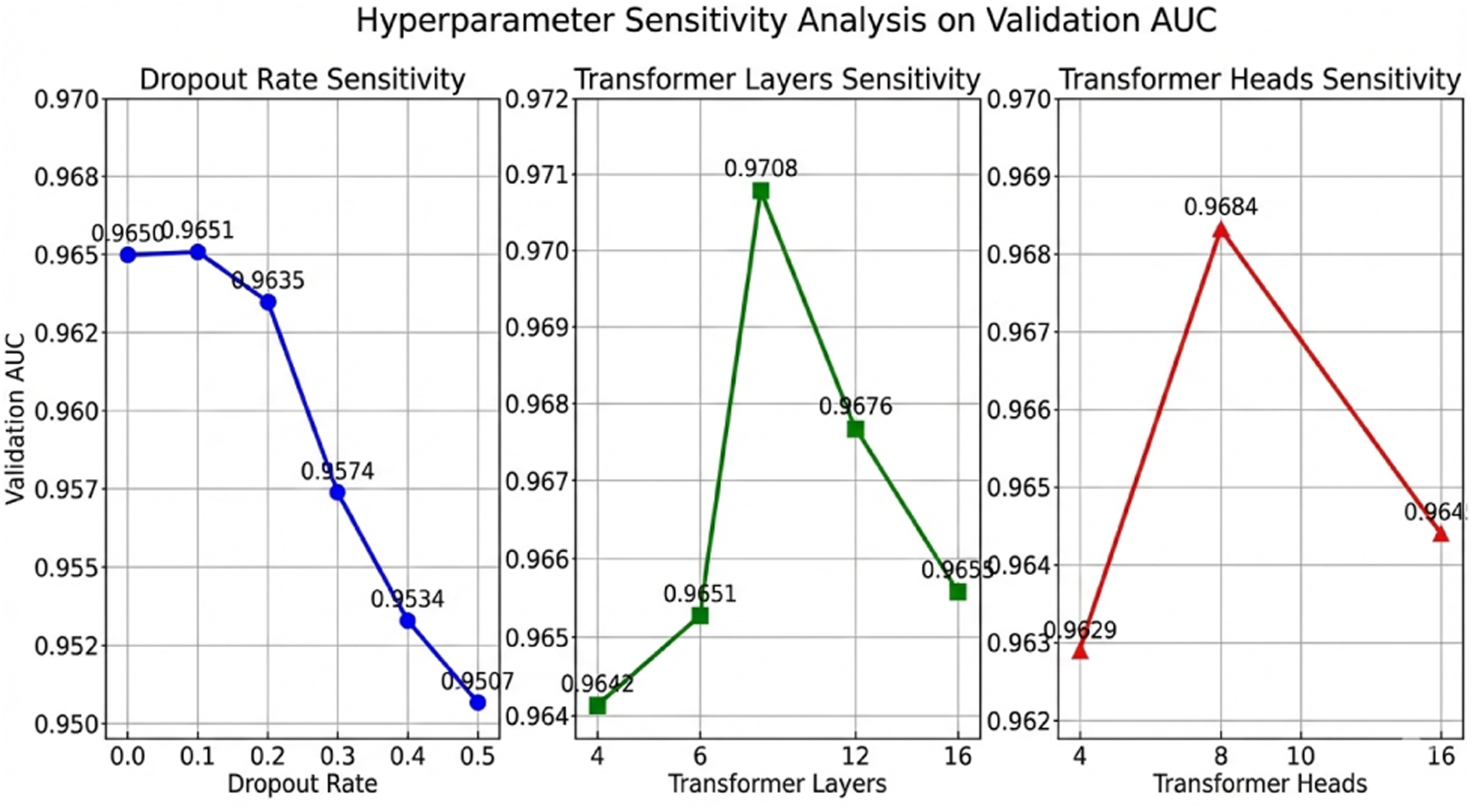
Hyperparameter Sensitivity Analysis.

#### 2.5.3. Ablation Study

To strictly evaluate the contribution of the Transformer architecture and verify the optimal fusion strategy for multi-view ultrasound data, we implemented two comparative baseline models using the same EfficientNetV2 backbone.

- CNN-Only (Global Average Pooling Baseline): This baseline represents a bag-of-features approach that aggregates multi-view information without modeling inter-view relationships.

- Batch Processing: The input tensor of shape [*B*,*N*,*C*,*H*,*W*] is reshaped to [*B*□*N*,*C*,*H*,*W*] to treat all images across the batch as independent samples.
- Feature Extraction: The shared CNN backbone extracts high-dimensional features from these images.
- Reshaping & Masking: The features are reshaped back to [*B*,*N*,*D*] (where *D*=1280). A binary mask is applied to zero out features corresponding to padded dummy images, ensuring they do not affect the calculation.
- Aggregation: We apply Global Average Pooling along the view dimension for valid images only. This produces a single, averaged feature vector for the patient, which is fed into the classification head. This model assumes all views contribute equally and independently to the diagnosis.
- CNN-RNN (Sequential Modeling Baseline): This model tests the hypothesis that sequential dependencies (temporal logic) exist within the multi-view acquisition process.

- Sequential Modeling: Unlike the parallel processing of the Transformer or the simple averaging of the CNN-Only model, this architecture utilizes a Long Short-Term Memory (LSTM) network.
- Temporal Dependencies: The sequence of image features extracted by the CNN is fed into the LSTM, which updates its hidden state step-by-step, attempting to capture temporal correlations between successive ultrasound frames.
- Final Representation: Instead of using all states, we extract the hidden state of the last valid time step (determined by the sequence length of non-padded images) to serve as the comprehensive representation of the lesion. This vector is then passed to the classifier to output the final prediction.

#### 2.5.4. State-of-the-Art Benchmarks

To validate the competitiveness of our proposed method against established solutions in medical image analysis, we benchmarked our model against three advanced architectures representing different fusion paradigms, all comparative models were retrained on our training set using their official implementations and optimal hyperparameters recommended in their original papers:

- VGG-ViT: A classic hybrid architecture that combines a VGG convolutional backbone with a Vision Transformer (ViT). This model serves as a benchmark for early CNN-Transformer integration strategies.
- TransMed: Originally designed for multi-modal medical image fusion, this model was adapted to process multi-view single-modality data. It serves to compare our method against complex, general-purpose medical transformers.
- MedFormer: A specialized Transformer architecture optimized for medical imaging tasks.

Comparing against MedFormer allows us to assess whether our lightweight EfficientNetV2-Transformer design can compete with or outperform purpose-built medical transformers in specific breast ultrasound applications.

### 2.6. Experimental Settings and Training Parameters

All experiments were conducted in the following environment:

- **Hardware Platform:** NVIDIA GeForce RTX 4060Ti GPU
- **Software Environment:** Python 3.10, PyTorch 2.7.1, torchvision, scikit-learn, pandas, numpy

To ensure the reproducibility of the experiments, the detailed hyperparameters are provided in Table 3.

**Table 3:** Hyperparameter configuration for model training.

| Parameter | Baseline Model | Hybrid Model |
| --- | --- | --- |
| Optimizer | AdamW | AdamW |
| Initial Learning Rate | $1 \times 10^{-4}$ | $5 \times 10^{-5}$ |
| Learning Rate Scheduler | Cosine Annealing | Cosine Annealing |
| Batch Size | 32 (images) | 4 (lesions) |
| Epochs | 100 (with Early Stopping) | 100 (with Early Stopping) |
| Early Stopping Patience | 5 | 5 |
| Monitored Metric | Validation AUC | Validation AUC |
| Loss Function | Binary Cross-Entropy | Binary Cross-Entropy |

### 2.7. Evaluation Metrics

Model performance was assessed using five standard binary classification metrics.

- Accuracy: Measures the proportion of all predictions that are correct.

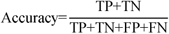
- Sensitivity: Measures the model’s ability to correctly identify malignant samples.

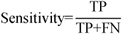
- Specificity: Measures the model’s ability to correctly identify benign samples.

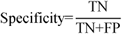

where TP, TN, FP, and FN represent correctly classified malignant lesions, correctly classified benign lesions, benign lesions misclassified as malignant, and malignant lesions misclassified as benign, respectively.
- **AUC :** Measures the model’s overall discriminative ability across all classification thresholds. An AUC closer to 1 indicates better performance.

### 2.8. Evaluation Metrics and Statistical Analysis

Performance was assessed using Accuracy, Sensitivity, Specificity, and AUC. Statistical Rigor:

- **Confidence Intervals (CI):** We calculated 95% CIs for all metrics using bootstrapping (n=1000 resamples) to demonstrate the stability of our results.
- **DeLong Test:** Used to statistically compare the AUC of different models. A p-value <0.05 was considered significant.

## 3. Results

### 3.1. Overall Model Performance Comparison

The proposed hybrid CNN-Transformer demonstrated a significant improvement in diagnostic performance over the conventional single-image baseline model across all evaluation metrics. This advantage was consistent on both the internal test set and, crucially, on the independent external dataset, underscoring the model’s robustness and generalization capabilities.

On the internal test set, which was partitioned at the tumor level to prevent data leakage, the hybrid model achieved an accuracy of 0.960 and an AUC of 0.9788. When challenged with the external dataset, comprising data acquired with different equipment, the model’s performance remained remarkably stable, yielding an accuracy of 0.940 and an AUC of 0.9730. In contrast, the baseline EfficientNetV2 model, which classifies each image independently, achieved lower metrics on both test sets. These results, detailed in Table 4, provide strong evidence that the fusion of multi-view information through the Transformer architecture leads to a significant leap in diagnostic power.

**Table 4:** Performance comparison of the two models on the test sets.

| Model | Test Set | Accuracy | Sensitivity | Specificity | AUC |
| --- | --- | --- | --- | --- | --- |
| Baseline Model | Internal | 0.881 | 0.863 | 0.914 | 0.9521 |
|  | External | 0.894 | 0.909 | 0.818 | 0.9437 |
| **CNN-Transformer** | Internal | 0.960 | 0.967 | 0.954 | 0.9788 |
|  | External | 0.940 | 0.952 | 0.929 | 0.9730 |

### 3.2. Analysis of Generalization Ability

To assess the model’s learning dynamics and potential for overfitting, the AUC was tracked for both the training and validation sets throughout the training process in Fig 5. The training and validation AUC curves rose in tandem and remained closely aligned, ultimately converging at a high level of performance (AUC>0.97). This indicates that the model did not overfit to the training data; instead, it learned generalizable features applicable to unseen data.

**Fig 5.**
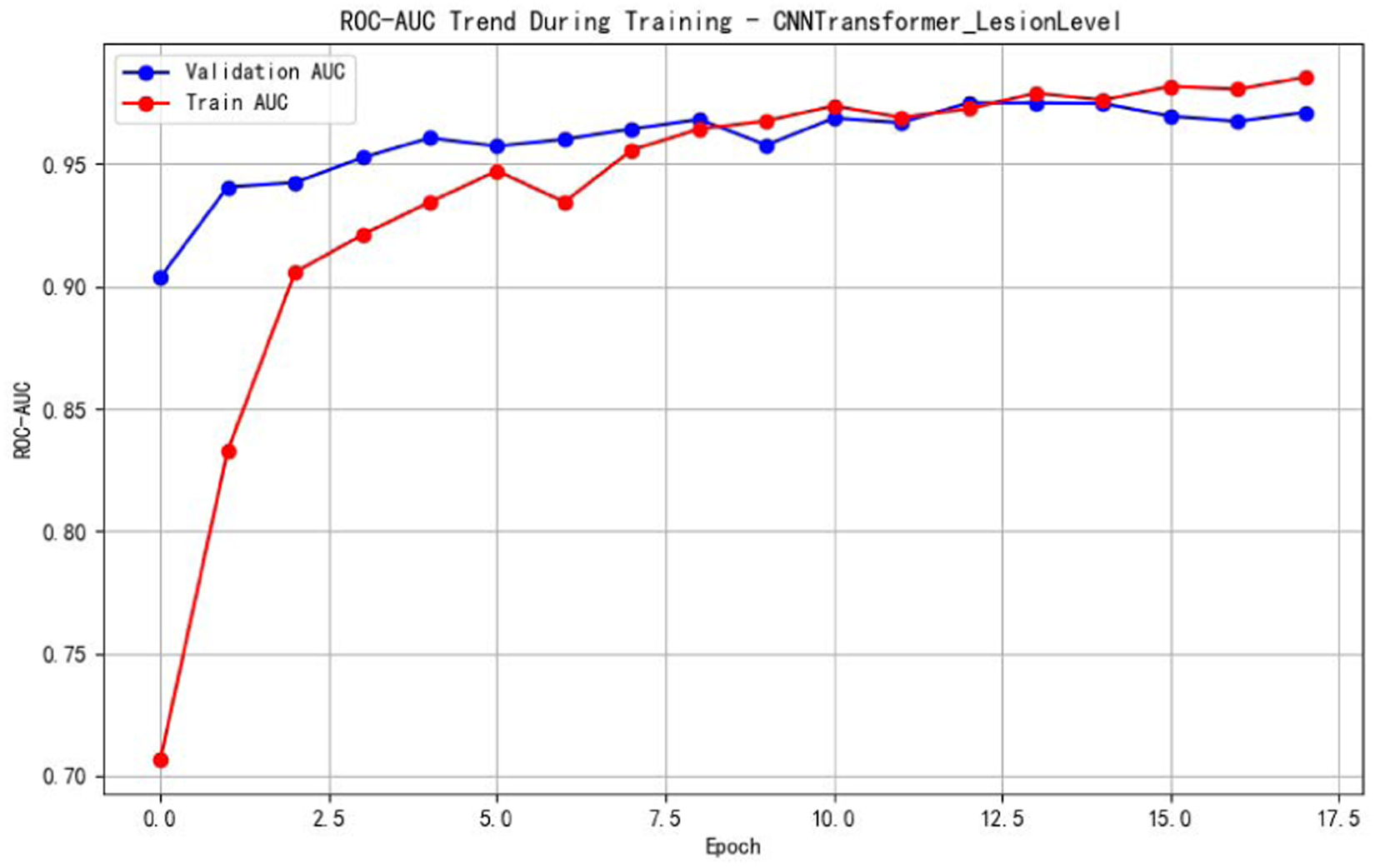
Model Training and Validation Curves (AUC vs. Epoch).

The ultimate test of generalization is performance on entirely new data. The Receiver Operating Characteristic (ROC) curves for the validation, internal test, and external dataset are presented in Fig 6. The curves are nearly superimposed, with high AUC values of 0.9708, 0.9788, and 0.9730, respectively. This significant consistency across three independent datasets strongly suggests that the model has learned robust, broadly applicable diagnostic features, validating both the architectural design and the rigorous data partitioning strategy.

**Fig 6.**
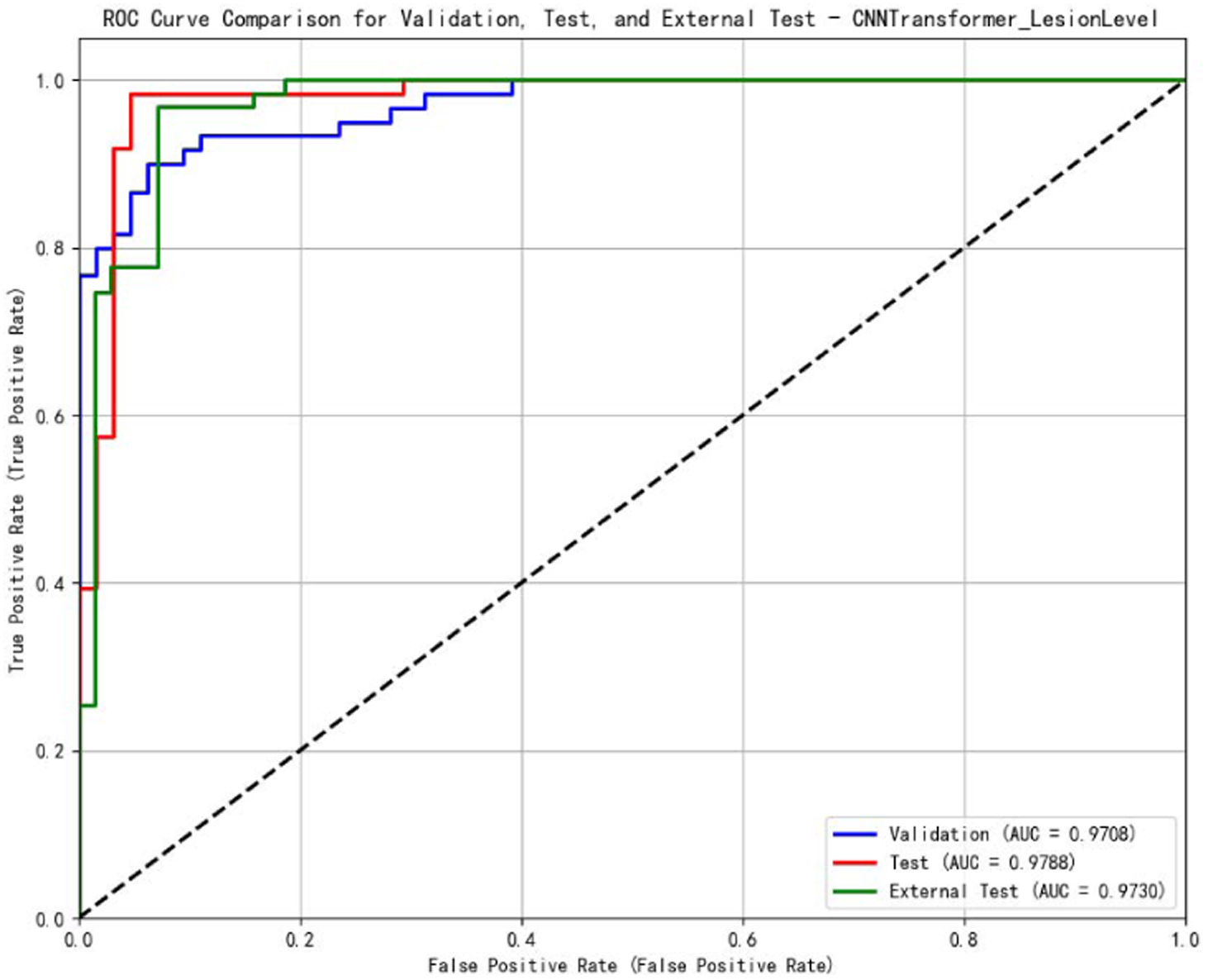
ROC Curves on Validation, Internal Test, and External Dataset.

To address the concern that deep learning models may overfit to specific device artifacts (e.g., speckle patterns specific to a vendor) and to ensure clinical generalization, we evaluated the model’s performance stratified by ultrasound machine manufacturer. The internal dataset was acquired using two distinct high-end systems: GE LOGIQ E9 and Philips EPIQ 7C, which employ different beamforming and post-processing algorithms. The model achieved an AUC of 0.9875 on the GE system and 0.9722 on the Philips system in Fig 7. The curves are nearly identical, and the high AUC across both vendors confirm that the model extracts robust, biologically relevant features rather than relying on equipment-specific noise or post-processing signatures. This minimal performance discrepancy (ΔAUC<0.02) demonstrates that the proposed architecture successfully mitigates the influence of machine heterogeneity, validating its potential for widespread deployment in multi-vendor clinical environments.

**Fig 7.**
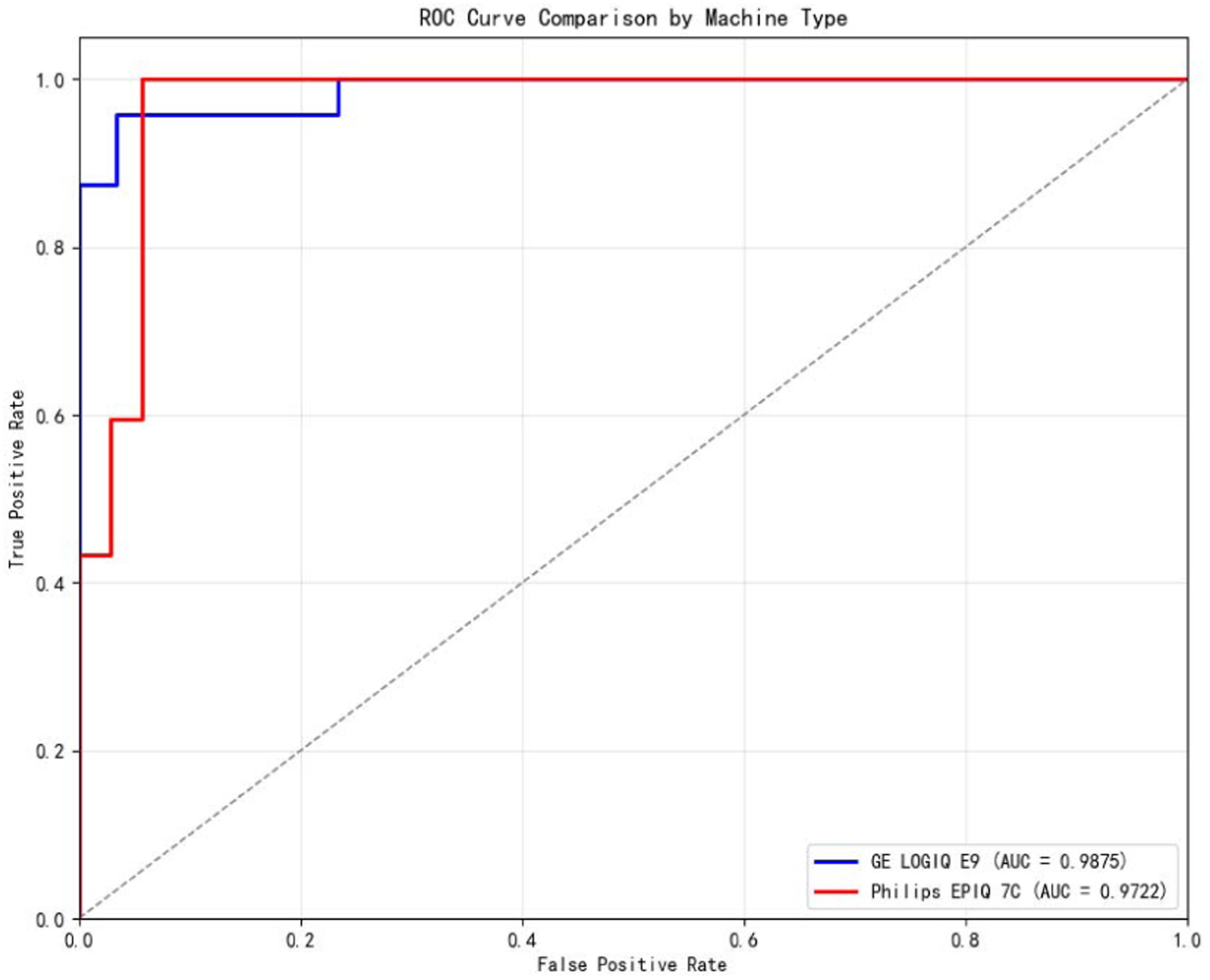
ROC Curves for both devices on Internal Test.

### 3.3. Detailed Classification Performance

A detailed breakdown of the model’s classification performance is presented via confusion matrices for the internal and external dataset (Fig 8) and a comprehensive classification report (Table 5).

**Fig 8.**
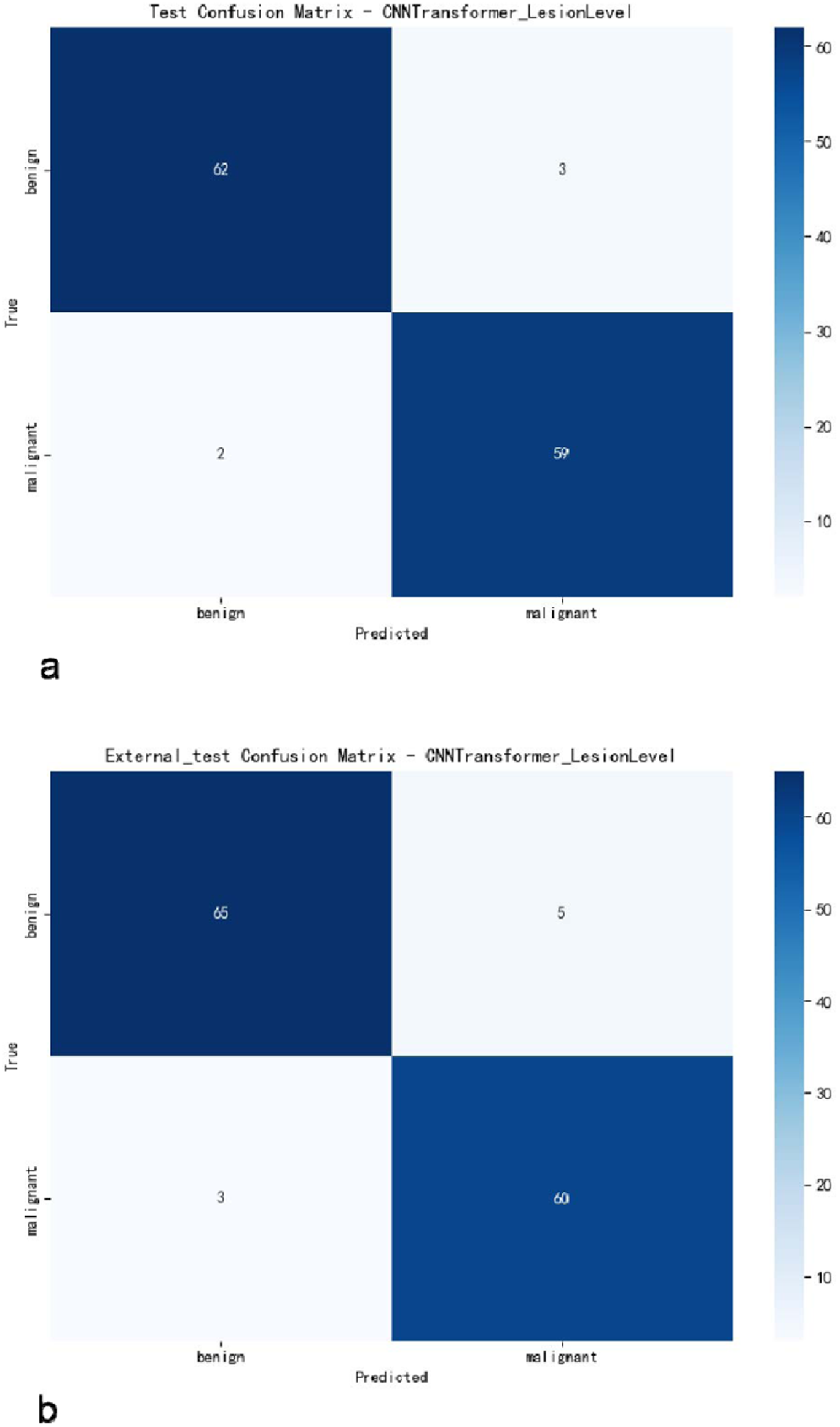
Confusion Matrices on the (a) Internal and (b) External Dataset.

**Table 5:** Detailed Classification Report on the Internal and External dataset.

| Test Set | Class | Precision | Recall | F1-Score | Support |
| --- | --- | --- | --- | --- | --- |
| Internal | Benign | 0.969 | 0.954 | 0.961 | 65 |
|  | Malignant | 0.952 | 0.967 | 0.959 | 61 |
|  | Macro Avg | 0.960 | 0.961 | 0.960 | 126 |
| External | Benign | 0.956 | 0.929 | 0.942 | 70 |
|  | Malignant | 0.923 | 0.952 | 0.938 | 63 |
|  | Macro Avg | 0.939 | 0.940 | 0.940 | 133 |

On the internal test set, the confusion matrix shows the model correctly identified 62 of 65 benign lesions and 59 of 61 malignant lesions in Fig 8a. As detailed in Table 5, this performance corresponds to a recall of 0.954 (specificity) and a precision of 0.969 for benign cases. For the crucial task of identifying malignancy, the model achieved a Sensitivity of 0.967 and a precision of 0.952. The excellent F1-scores of 0.960 for both classes highlight a strong and desirable balance between precision and recall.

The model’s robustness was further validated on the external dataset in Fig 8b. It achieved a recall (specificity) of 0.929 and a precision of 0.956 for benign lesions. For malignant lesions, it demonstrated an even higher recall (sensitivity) of 0.952, with a corresponding precision of 0.923. This indicates a very low rate of missed malignant cases. The resulting F1-scores of 0.942 for benign and 0.938 for malignant classes confirm the model’s strong diagnostic capability on unseen data.

In summary, the consistently high performance across both datasets, highlighted by macro-averaged F1-scores of 0.960 (internal) and 0.940 (external), affirms the model’s reliability and strong generalization potential.

### 3.4. Analysis of Prediction Probability and Confidence

Beyond standard classification metrics, the confidence of a model’s predictions is a critical dimension of its clinical reliability. Fig. 9 illustrates the frequency distribution of predicted malignancy probabilities for all benign and malignant samples in the test sets. The histograms reveal a distinct separation: probabilities for benign samples (blue) are highly concentrated near 0.0, while those for malignant samples (red) are predominantly clustered near 1.0. This distinct lack of overlap between the two distributions visually confirms the model’s strong discriminative ability. It demonstrates that the model does not merely achieve high accuracy but renders diagnostic decisions with high confidence.

**Fig 9:**
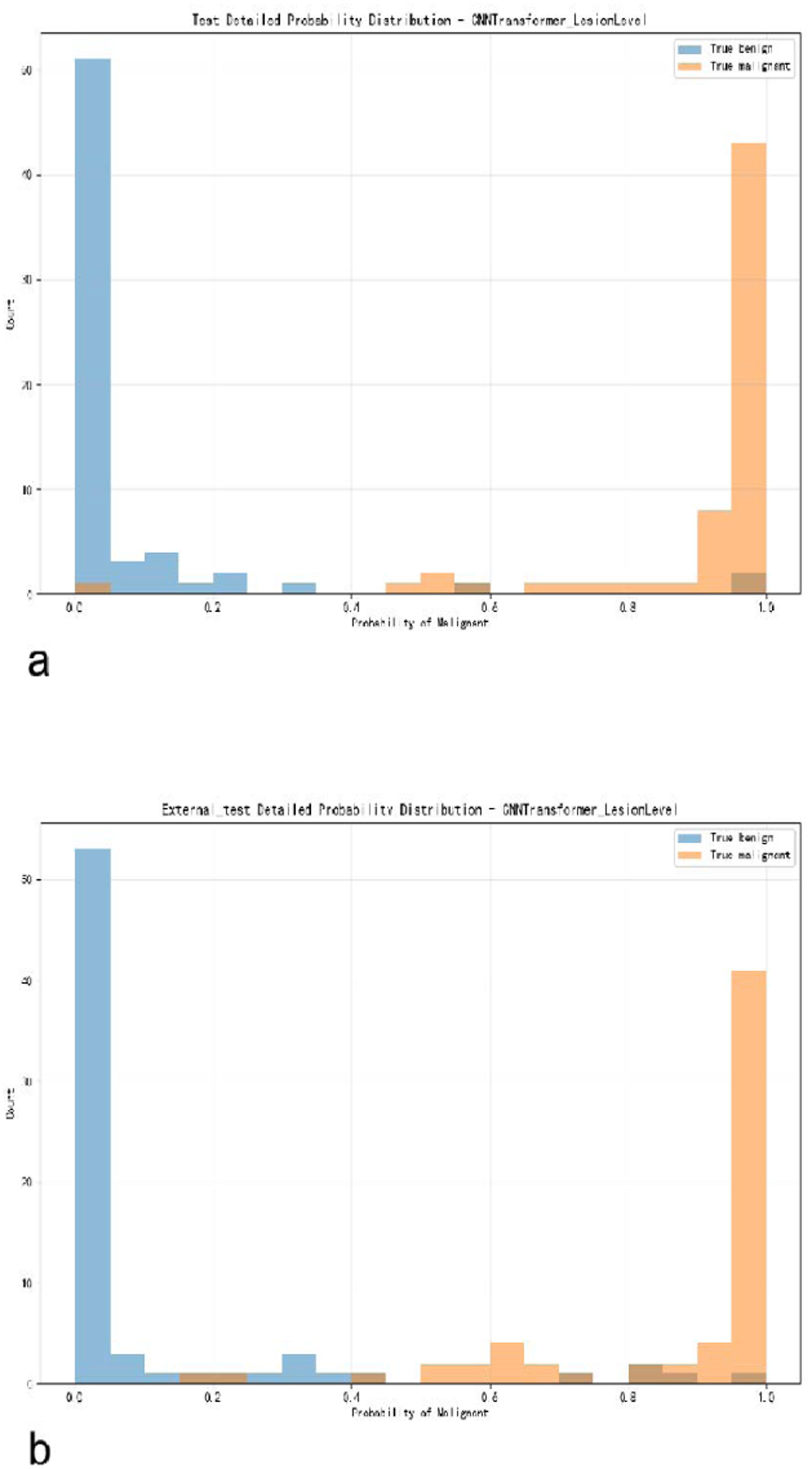
Histogram of predicted probability distributions on the (a) internal and (b) external dataset.

### 3.5. Confidence Intervals Analysis

The proposed hybrid CNN-Transformer demonstrated robust performance on both internal and external dataset. The results, including 95% Confidence Intervals, are presented in Table 6. The model achieved an AUC of 0.9788 on the internal set and maintained an AUC of 0.9730 on the external dataset. The overlap in the 95% CIs between internal and external performance indicates excellent generalization; the model did not simply memorize the training data distribution or specific hospital artifacts. The high Sensitivity (>0.95 in both sets) is clinically pivotal, ensuring that malignant cases are rarely missed.

**Table 6:** Confidence interval analysis results.

| Test Set | Accuracy | AUC | Sensitivity | Specificity |
| --- | --- | --- | --- | --- |
| Internal | 0.9603 | 0.9788(95%CI: 0.9508 - 0.9990) | 0.967(95%CI:0.917 - 1.0000) | 0.954(95%CI: 0.900 - 1.000) |
| External | 0.9398 | 0.9730(95%CI: 0.9414 - 0.9937) | 0.952(95%CI:0.894 - 1.000) | 0.929(95%CI: 0.864 - 0.985) |

### 3.6. Ablation Study Results

To determine the optimal mechanism for multi-view information integration, a comprehensive component-wise ablation study was conducted. The objective was to evaluate the hypothesis that modeling ultrasound views as an unordered set—via a Transformer-based architecture—yields superior performance compared to sequential modeling using RNNs or view-independent processing with CNNs alone (Table 7).

**Table 7:** Ablation study results.

| Model Architecture | Fusion Strategy | Validation AUC | Test AUC |
| --- | --- | --- | --- |
| CNN-Only | Max Pooling (Post-hoc) | 0.9502 | 0.9576 |
| CNN-RNN | Bi-Directional LSTM | 0.9676 | 0.9659 |
| **CNN-Transformer** | Self-Attention (Proposed) | **0.9708** | **0.9788** |

The CNN-Only model achieved an AUC of 0.9576, establishing a competitive baseline; however, it is limited to post-hoc aggregation of view-specific features without cross-view feature synthesis. The CNN-RNN model (AUC 0.9659) introduces feature-level fusion through sequential modeling, yet it is outperformed by the CNN-Transformer model (AUC 0.9788). This statistically significant improvement supports our hypothesis: the permutation invariance inherent in the Transformer architecture is more suitable for multi-view ultrasound analysis than the ordered assumption underlying RNNs. By employing self-attention, the Transformer dynamically prioritizes the most informative views irrespective of their positional order, thereby emulating a cognitive integration process free from artificial temporal constraints.

### 3.7. Computational Efficiency

To assess clinical feasibility, our hybrid CNN-Transformer measured inference time on an NVIDIA RTX 4060Ti.

- Total Time (126 samples): 4.2249 seconds.
- Average Time per Patient: 0.0335 seconds.

This ultra-low latency (<40ms) confirms the model is suitable for real-time integration into clinical workflows.

### 3.8. Comparison with State-of-the-Art Architectures

To strictly validate the competitiveness of our proposed method, we benchmarked it against three advanced architectures representing different fusion paradigms: VGG-ViT (classic hybrid), TransMed (multi-modal fusion), and MedFormer (specialized medical transformer). As detailed in Table 8, our EfficientNetV2-Transformer hybrid model achieved the highest Test AUC of 0.9788, outperforming all comparative baselines. A deeper analysis reveals the architectural reasons behind this performance gap:

**Table 8:**
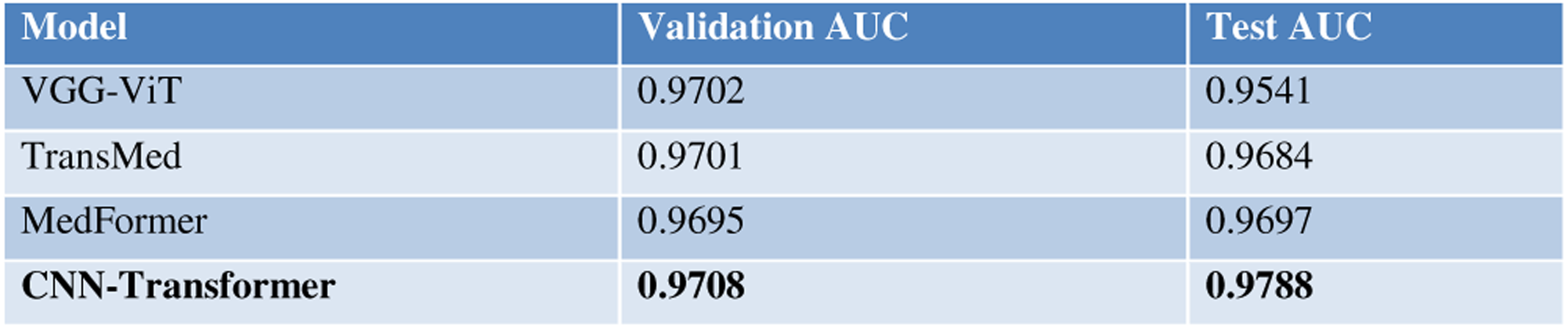
Comparison results of similar models.

#### 3.8.1. Superior Generalization vs. VGG-ViT

The VGG-ViT model exhibited a high Validation performance of AUC with 0.9702, but its performance dropped sharply on the Test set (0.9541). This suggests that the heavy parameter count of the VGG backbone, combined with a standard Vision Transformer, led to overfitting on the ultrasound dataset. In stark contrast, our model achieved superior generalization, improving from a Validation AUC of 0.9702 to a Test AUC of 0.9788. This confirms that our choice of the lightweight EfficientNetV2 backbone combined with a shallow (8-layer) Transformer strikes a better balance between model complexity and data efficiency, learning robust pathological features rather than memorizing noise.

#### 3.8.2. Task-Specific Optimization vs. TransMed and MedFormer

TransMed (Test AUC 0.9684) and MedFormer (Test AUC 0.9697) represent complex, heavy-weight architectures originally designed for multi-modal tasks (e.g., MRI-CT fusion) or general medical segmentation. While they performed well, they did not surpass our proposed model.

- The Bag-of-Images Advantage: TransMed assumes complex inter-modal relationships that are unnecessary for single-modality ultrasound. Our model, by removing positional encoding, enforces permutation invariance. This design is specifically optimized for the unordered nature of multi-view ultrasound (where the order of images does not matter), allowing the model to focus purely on aggregating complementary diagnostic evidence.
- Efficiency: Our model achieves superior accuracy without the computational burden of these heavy transformers. This demonstrates that a focused, task-specific architecture is more effective for breast ultrasound classification than adapting large-scale, general-purpose medical transformers.

In summary, the benchmarking results demonstrate that increased model complexity does not necessarily translate to superior diagnostic performance. Our model’s success stems from aligning the network architecture with the clinical reality of breast ultrasound—treating views as an unordered set and prioritizing feature robustness over model complexity.

### 3.9. Model Interpretability using Gradient-weighted Class Activation Mapping (Grad-CAM)

To investigate the model’s decision-making basis, Grad-CAM was used to visualize the regions of interest the CNN feature extractor focused on clinical ultrasound features. Although the final diagnosis results from the Transformer’s fusion of features, Grad-CAM provides insight into whether the foundational feature extraction is clinically relevant.

As shown in Fig 10 and Fig 11, when the model correctly classifies a lesion, the high-attention areas (heatmaps) precisely overlap with clinically significant pathological features, such as irregular margins, internal microcalcifications, and angular contours. This visualization confirms that the CNN component effectively learns to identify meaningful biomarkers, providing a reliable and interpretable foundation for the Transformer’s subsequent global analysis. This enhances trust in the model’s decisions by demonstrating that its reasoning is aligned with established clinical diagnostic criteria.

**Fig 10.**
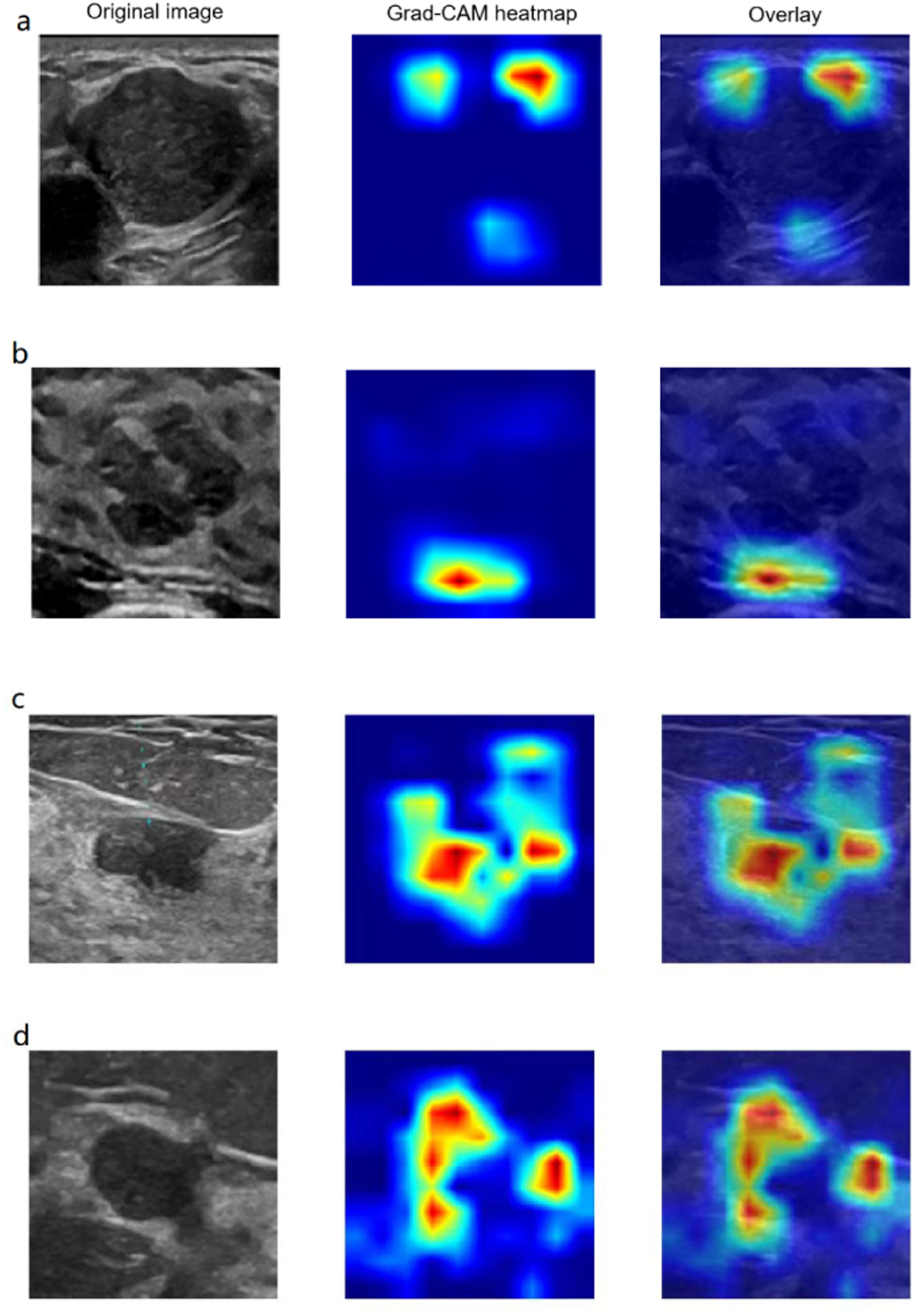
BI-RADS 4a Breast Lesion with Benign Features on Ultrasound and Grad-CAM Analysis. Ultrasound imaging reveals a solid, hypoechoic mass classified as BI-RADS 4a. The Hybrid model correctly predicted this lesion as benign. Corresponding Grad-CAM overlays visually explain the model’s decision by highlighting key benign features: (a) obtuse marginal angles, (b) significant posterior acoustic enhancement, (c) homogeneous internal echotexture, and (d) a well-circumscribed boundary.

**Fig 11.**
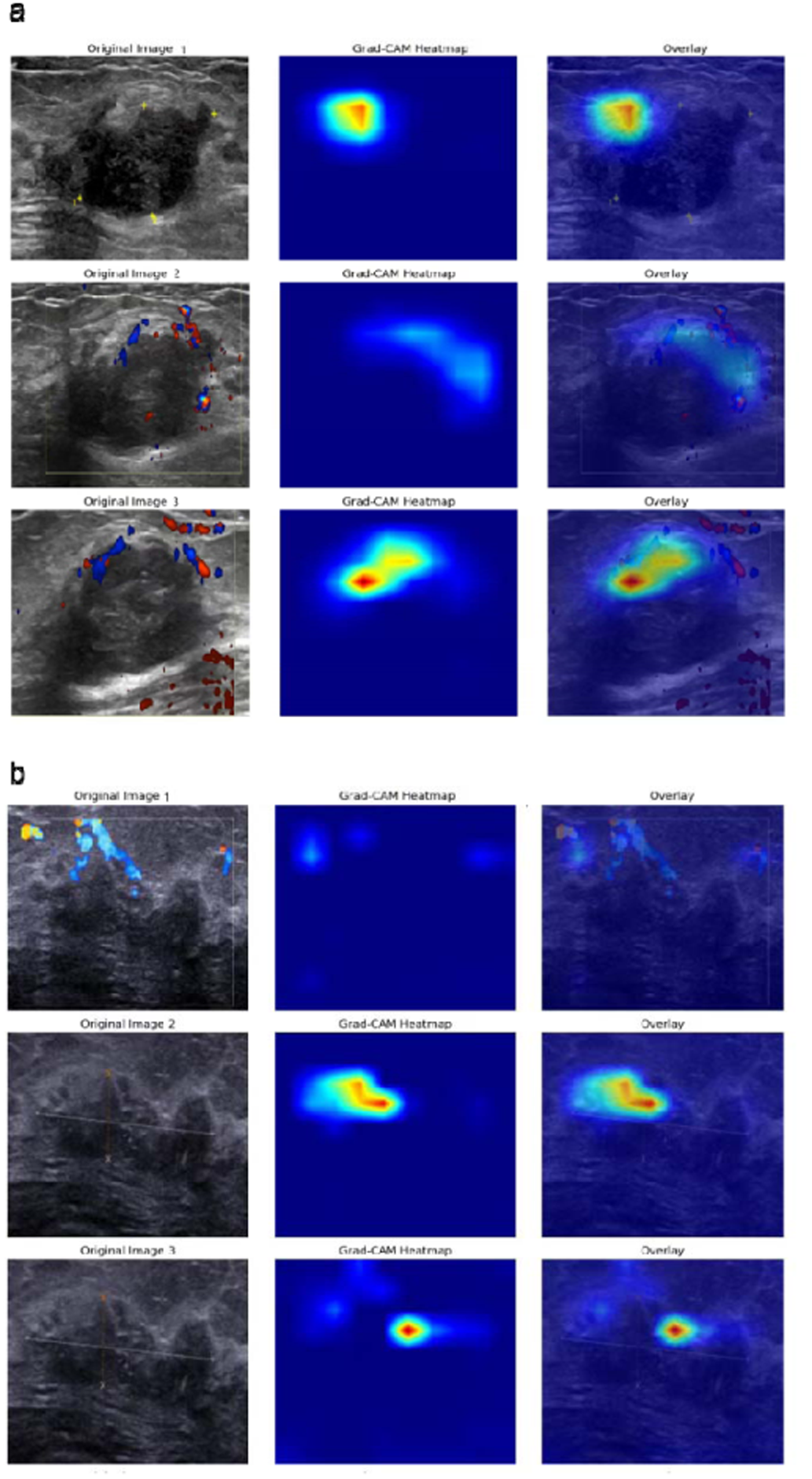
BI-RADS 4c Breast Lesion with Malignant Features on Ultrasound and Grad-CAM Analysis. Ultrasound imaging depicts a mass with highly suspicious characteristics, classified as BI-RADS 4c. The Hybrid model accurately predicted this lesion as malignant. The explanatory Grad-CAM overlays identify the critical malignant features driving the model’s decision: (a) angular, spiculated margins, and rich internal vascularity on color Doppler imaging, (b) a thick echogenic halo, an irregular and indistinct boundary.

## 4. Expert Validation Test

To validate the clinical relevance of our model and address concerns regarding human-machine comparison, we conducted a blind test using the internal test set (n=126). Three ultrasound physicians participated: two junior doctors (3-5 years of experience) and one senior doctor (>15 years of experience). They independently classified the 126 lesions without access to the model’s predictions or pathology results (Table 9).

**Table 9:** Comparison with Human Experts.

| Method | Correct Cases | Accuracy | Sensitivity | Specificity |
| --- | --- | --- | --- | --- |
| Junior Doctor 1 | 103 | 0.818 | 0.820 | 0.815 |
| Junior Doctor 2 | 104 | 0.825 | 0.803 | 0.846 |
| Senior Doctor | 107 | 0.849 | 0.869 | 0.831 |
| Average Human | 104.7 | 0.831 | 0.831 | 0.831 |

As presented in Table 9, the proposed model significantly outperformed all human readers. The model achieved an accuracy of 0.960, surpassing the senior expert (0.849) by over 11 percentage points. Notably, the model’s sensitivity (0.967) was substantially higher than even the senior doctor (0.8689), indicating a superior capability in detecting malignancies and reducing false negatives. This suggests that the model can serve as a highly effective suggestion to reduce missed diagnoses in clinical practice.

To understand the source of this performance gap, we analyzed discordant cases where human experts erred but the model succeeded. Fig 12 illustrates a challenging diagnostic scenario involving a hypoechoic mass with ambiguous features. The original grayscale image (Fig 12a) and Color Doppler (Fig 12b) show a hypoechoic mass with somewhat ambiguous features, possibly exhibiting minor irregularities or shadowing that mimics malignancy, leading to a potential human misdiagnosis (False Positive).

- Grad-CAM Insight: The heatmap reveals that the model’s attention was not distracted by the internal heterogeneity. Instead, it focused strongly on the upper and lateral boundaries of the lesion. By accurately identifying the continuity of the tumor and the distinct tissue interface (highlighted in red), the model confirmed the well-circumscribed nature of the tumor.
- Clinical Significance: This case exemplifies the model’s robustness in reducing unnecessary biopsies. While human eyes might be misled by suspicious-looking artifacts, the Grad-CAM confirms that the model grounds its decision on subtle but definitive markers of benignity, effectively serving as a gatekeeper to improve specificity.

**Fig 12.**
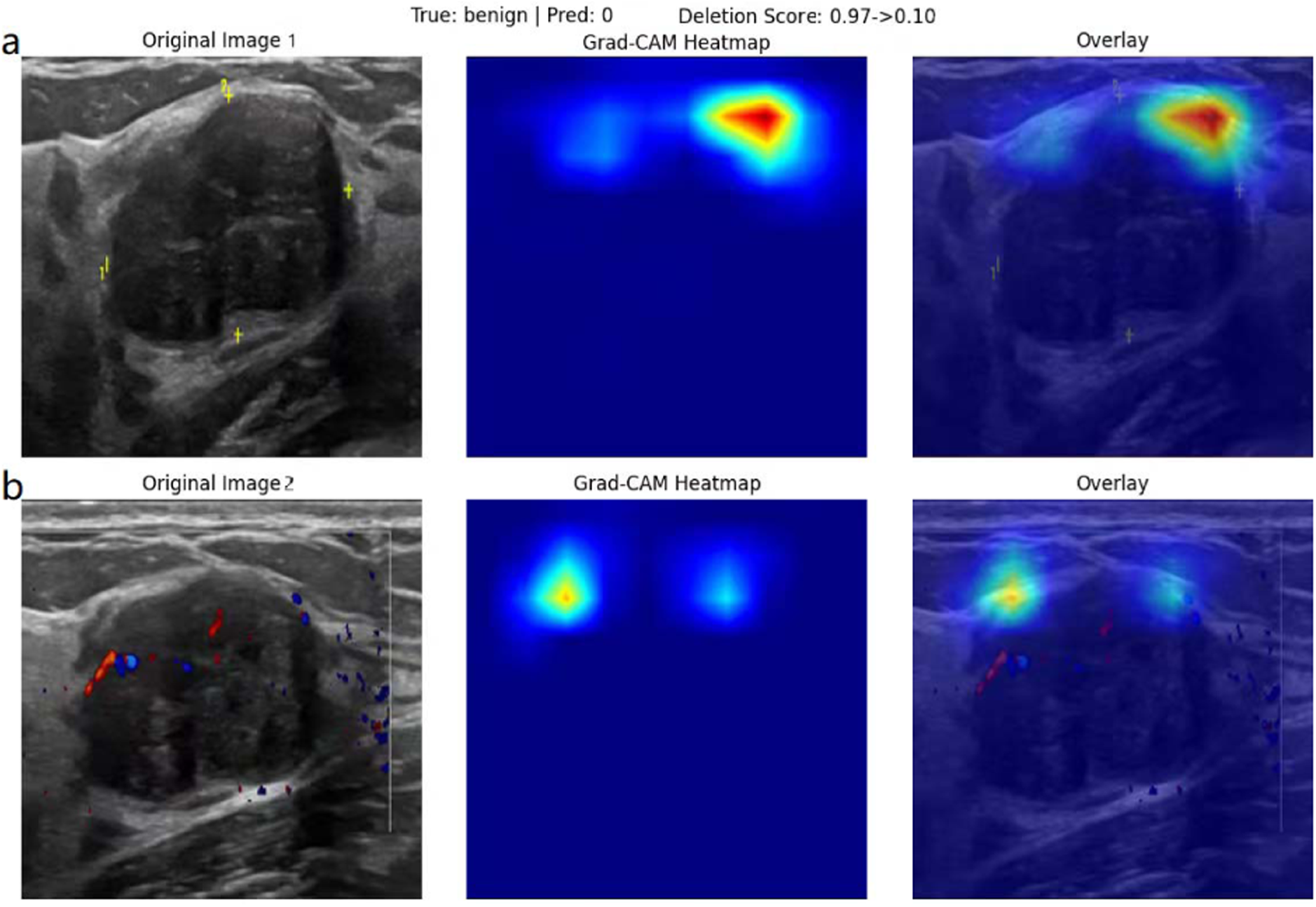
A Challenging Case Correctly Reclassified by the Hybrid Model (Human-AI Discordance). (a) B-mode ultrasound and (b)Color Doppler ultrasound, where the Grad-CAM heatmap focused strongly on the upper and lateral boundaries of the lesion. It identifies the continuity of the tumor and the distinct tissue interface (highlighted in red).

## 5. Prospective Validation on a Newly Collected Dataset

### 5.1. Rationale and Data Collection and Inclusion

To rigorously address potential concerns regarding model generalization and data leakage, we performed a stringent prospective validation of the final hybrid CNN-Transformer. This test used a completely new, independent dataset collected after the development and finalization of the model, mimicking a real-world clinical deployment scenario.

Data for this prospective test were collected consecutively from September 2025 to December 2025. A total of 188 new lesions (including 122 benign and 66 malignant cases, confirmed by pathology) were collected during this period, ensuring no overlap with the initial training, validation, or internal test sets. All data collection and image acquisition protocols remained identical to the procedures outlined in Section 2.

### 5.2. Prospective Test Results

The hybrid CNN-Transformer was applied to the newly collected prospective test set. The results, visualized by the Classification Report, Confusion Matrix, and ROC Curve (Fig 13), demonstrate the model’s robust generalization capability.

**Fig 13.**
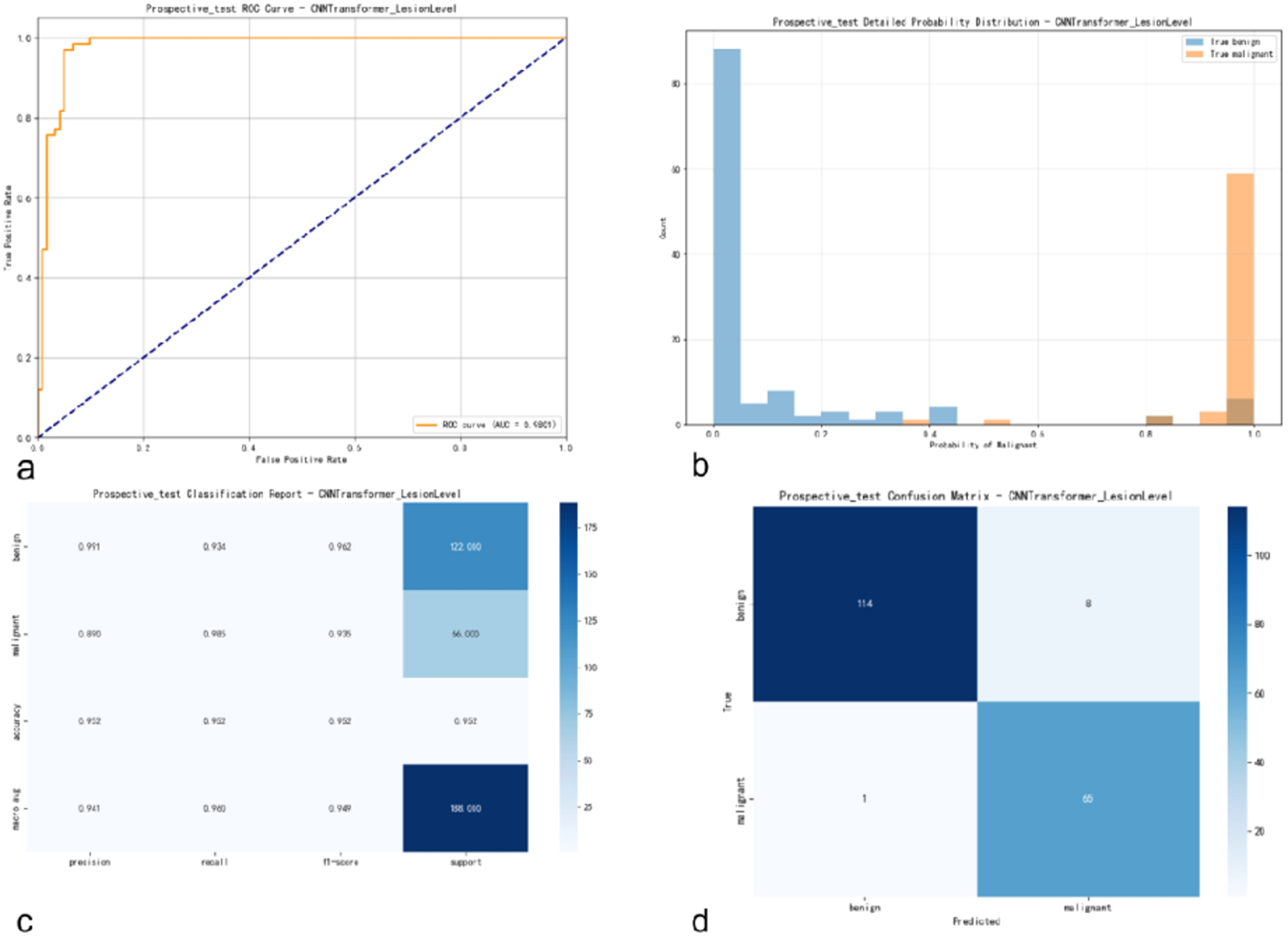
Performance Evaluation on the Prospective Validation Cohort. (a) ROC Curve: The model achieved an impressive AUC of 0.9801 on the prospective dataset. (b) Detailed Probability Distribution: The histogram shows a clear separation between predicted probabilities for benign (blue) and malignant (orange) cases, indicating high prediction confidence. (c) Classification Report: Quantitative metrics detailing the Precision, Recall, and F1-score for each class, with an overall accuracy of 95.2%. (d) Confusion Matrix: The matrix confirms 98.48% sensitivity (65/66) for malignant lesions and highlights that 8 benign cases were misclassified (114/122 correctly identified).

The model achieved an overall accuracy of 95.2% and AUC of 98.01%.Specifically, the model demonstrated a strong capability to correctly identify 65 of 66 malignant lesions and 114 of 122 benign lesions.

These results are comparable to the performance observed on the internal test set (Section 3.1), confirming that the multi-view CNN-Transformer architecture is highly effective and stable across distinct, temporally separated, real-world clinical data.

## 6. Discussion

The results of this study indicate that the proposed hybrid CNN-Transformer is significantly superior to conventional single-image CNN approaches for classifying breast tumors in ultrasound images. The core reason for this performance gain lies in an architecture that more faithfully emulates the cognitive process of clinical experts.

The effectiveness of multi-view fusion is the central finding. A single-image model processes each view in isolation, whereas the Transformer encoder in our hybrid model functions as a virtual expert panel. Its self-attention mechanism allows the model to dynamically weigh and integrate information from the entire set of images for a given lesion(21, 22). For instance, it can learn to assign greater importance to a view that clearly depicts spiculated margins while down-weighting another view obscured by acoustic artifacts. This dynamic, global evidence integration is a powerful analogy for expert diagnostic reasoning and is the key driver behind the model’s performance leap. This architecture also confers greater robustness to the variations inherent in real-world clinical data, such as differences in image quality or the number of available views, as the model can learn to prioritize high-quality, informative images(23, 24).

The clinical implications of the model’s performance are substantial. The high specificity achieved on both internal (0.954) and external (0.929) dataset indicates a strong ability to correctly identify benign lesions. This could translate to a significant reduction in false-positive findings, potentially sparing many women from the anxiety, physical discomfort, and cost of unnecessary biopsies. Concurrently, the model’s high sensitivity (0.967 internal, 0.952 external) facilitates a low rate of missed malignant tumors, which is critical for facilitating early diagnosis and treatment.

Despite its promising results, this study has limitations that outline directions for future research. First, although the model demonstrated robust generalization on an independent external dataset and across two different ultrasound vendors (GE and Philips). To fully establish the model’s reliability in diverse clinical settings, large-scale multi-center validation encompassing a wider array of equipment manufacturers and patient demographics is necessary. Second, while our Grad-CAM analysis confirmed that the model focuses on clinically relevant features and the expert validation study demonstrated its superiority over human readers, the current system provides only visual heatmaps and probability scores. It does not yet generate structured text explanations or Breast Imaging-Reporting and Data System(BI-RADS) descriptors, which would further enhance trust and facilitate seamless integration into the radiologist’s reporting workflow. Finally, although our prospective validation verified the model’s stability on real-world data, a long-term randomized clinical trial (RCT) is required to quantitatively assess its actual impact on reducing unnecessary biopsies and improving clinical workflow efficiency. By using a shallow Transformer (8 layers) on top of an efficient backbone (EfficientNetV2), we achieve a sweet spot between depth and speed. This model does not require a supercomputer; it can run on a standard medical workstation equipped with a mid-range GPU.

## 7. Conclusion

This study successfully developed and validated a novel hybrid CNN-Transformer deep learning model for breast ultrasound diagnosis. By synergistically combining the powerful local feature extraction of a CNN with the sophisticated global information fusion capabilities of a Transformer, the model effectively simulates the multi-view diagnostic process of clinical experts. Using a rigorous, tumor-level evaluation methodology, the proposed model demonstrated outstanding classification performance on an internal test set (AUC 0.9788) and proved its excellent generalization on an independent external dataset (AUC 0.9730). The consistently high accuracy, sensitivity, and specificity across diverse datasets significantly exceed those of traditional single-image methods. This work not only validates a superior technical architecture but also underscores the methodological importance of multi-view analysis and robust external validation. The model shows potential as a reliable tool to assist clinicians in making more accurate diagnoses of breast cancer, with the ultimate aim of improving patient health and well-being.

In future work, we will focus on developing a large scale study with more external validations. Ultimately, the goal is to integrate this technology into a user-friendly CAD system and conduct prospective clinical trials to evaluate its real-world impact on diagnostic accuracy, workflow efficiency, and patient outcomes.

## Footnote

## Ethical Statement

The authors are accountable for all aspects of the work in ensuring that questions related to the accuracy or integrity of any part of the work are appropriately investigated and resolved. This study was conducted in accordance with the Declaration of Helsinki (as revised in 2013). The medical ethics committee of the Second Affiliated Hospital of Guangzhou University of Chinese Medicine approved the study (Approval No. ZE2023-427).The requirement for informed consent was waived due to the retrospective nature of the study.

## Data Availability

The data that support the findings of this study are available from the corresponding author upon reasonable request.

